# Individual-and Community-Level Determinants of Zero-Dose Children in Nigeria: A Multilevel Analysis using the 2024 Nigerian Demographic and Health Survey

**DOI:** 10.64898/2026.04.18.26351159

**Authors:** Desalegn Mitiku Kidie, Abraham Dessie Gessesse, Tsegaamlak Kumelachew Derse, Tadios Lidetu, Addisu Simachew Asgai, Jenberu Mekurianew Kelkay

**Author notes:** **Author emails**, DM, AD, TK, TL, AS, JM. **Corresponding Author**: DM-*****Department of Pediatrics and Child Health Nursing, College of Health Science, Debark University, Debark, Ethiopia, <u>E-mail:</u>.

## Abstract

**Background:** Zero-dose children, defined as those who have not received the first dose of a diphtheria-tetanus-pertussis-containing vaccine (DPT1), are a key indicator of inequitable access to immunization services. Nigeria remains one of the largest contributors to the global burden of zero-dose children. This study estimated the prevalence of zero-dose children aged 12-23 months and identified individual-and community-level determinants using the 2024 Nigeria Demographic Health Survey (NDHS).

**Methods:** A secondary analysis of cross-sectional analysis was conducted using data from 4,711 children aged 12-23 months in the 2024 NDHS kids recode dataset. A multilevel mixed-effects logistic regression model was fitted to account for the hierarchical structure of the data. Four models were compared: null, individual-level, community-level, and combined models. Adjusted odds ratios (AORs) with 95% confidence interval (CI) were used to identify significant determinants at p<0.05.

**Results:** The weighted prevalence of zero-dose children was 37.3% (95% CI: 35.1-39.6%). Significant factors included birth order, maternal age, maternal occupation, parental education, household wealth, antenatal attendance, postnatal care utilization, place of delivery, religion, distance to health facilities, and geographical region. Children whose mothers had higher educational attainment, attending antenatal care, deliver in the health facilities, and received postnatal care were significantly less likely to be zero-dose status. Conversely, children from poorer households, those facing distance barriers to health facilities, those belongings to Muslim and traditional religion group and those residing in certain geographical regions had higher odds of zero-dose children, with significant regional variations observed.

**Conclusion:** zero-dose vaccination remains highly prevalent in Nigeria and is strongly influenced by socioeconomic disadvantage, maternal healthcare utilization, religion, and regional inequities. Strengthening integrated maternal and child health services and improving access in underserved regions are essential to achieving equitable vaccination coverage.

## Introduction

Immunization is a major driver of public health, protecting against a wide range of infectious diseases that contribute substantially to global morbidity and mortality(1–3).The Expanded Programme on Immunization (EPI), established by the world health organization (WHO) in 1974, has significantly increased global access to life-saving vaccines for children(4). Immunization defined as the administration of vaccine to stimulate the immune system and develop protection against specific disease(5), remains one of the most successful and cost-effective public health interventions, saving millions of live annually and improving global health outcomes (6).It is established that routine immunization will avert approximately 5.1 million child deaths worldwide between 2021 and 2030(7).

In response to persistent immunization challenges and inequitable access, the 73^rd^ World Health Assembly endorsed the Immunization Agenda 2030 (IA2030) in August 2020. The strategy envisions a world in which everyone, everywhere, and at every age fully benefits from vaccines for better health and well-being. To realize this vision, IA2030 set three 2030 goals: reducing vaccine preventable morbidity and mortality, improving equitable access to new and existing vaccines, and strengthening immunizations within primary health care to support universal health coverage and sustainable development(8). Progress is monitored through WHO and United Nations International Children’s Emergency Fund (UNICEF) coverage estimates with key targets of reducing by 50% the number of children missing the first dose of diphtheria-tetanus-pertussis-containing vaccine (DTPcv1) and achieving 90% coverage for the third dose (DTP cv3) by 2030 (8, 9).

However, despite notable progress in expanding childhood immunization coverage, substantial gaps remain. In 2024, an estimated 14.3 million children who did not receive even the first dose of diphtheria-tetanus-pertussis-containing vaccine, commonly referred to as “zero-dose“ children, reflecting only a slight improvement from 2023 and highlighting the persistent challenge of achieving IA2030 targets (8, 10).

In 2021, an estimated 18.2 million zero-dose children were reported globally, representing over 70% of all under-immunized children and emphasizing the need to reach these populations to achieve IA2030 and Sustainable Development Goal targets (11, 12). These children are disproportionately concentrated in low-and middle-income countries, especially in urban slums, remote rural areas, and conflict-affected settings, where limited health service access, socioeconomic inequities, and structural barriers sustain persistent immunization gaps (13–15).

Globally, pooled studies in 2022 estimated over 17 million zero-dose children, while recent DTP1 coverage as a proxy for vaccine access reached about 89% in 2024, leaving roughly 11% of infants partially or completely unvaccinated. Most zero-dose children are concentrated in low-and lower -middle-income countries, which account for 87% of global burden (8, 16, 17). A small group of high population countries carries a disproportionate share of it. In 2024, more than half of all zero-dose children were concentrated in nine countries, including Nigeria, India, Sudan, the Democratic Republic of Congo, Ethiopia, and Indonesia (18, 19).

A study across remote-rural, urban setting and conflict-affected setting in low-and middle-income countries reported that of the 14,030,486 children unvaccinated for DPT1, over 11% (1,656,757) were in remote-rural areas, more than 28% (2,849,671 and 1,129,915) in urban and peri-urban areas, and up to 60% in other settings (19). The prevalence of zero-dose in Somalia, approximately 60.2% of children were zero-dose(20). Zero-dose status was associated with rural residency, female sex, non-institutional delivery, fewer than four antenatal care (ANC) visits, lack of postnatal care, poor maternal health seeking behavior, low parental education, limited maternal empowerment and decision-making power, financial barriers, lack of media access, low community literacy, and low country-level health expenditure(15, 17, 20, 21).

Therefore, this study aimed to assess the prevalence of zero-dose immunization and identify its determinants using the recent Nigeria 2024 Demographic health survey (DHS) data. Understanding these factors is crucial to reducing the burden of zero-dose children and improving child health outcomes, particularly in preventing communicable and vaccine preventable disease. Specifically, the study examined the prevalence of zero-dos vaccination and the individual and community level determinants among children age 12-23 months through a cross sectional analysis of 2024 demographic health survey.

### Objective

To determine the prevalence of zero-dose children aged 12-23 months using 2024 Nigerian demographic health survey

To identify individual and community level determinant factors of zero-dose children age 12-23 months using 2024 Nigerian demographic health survey

## Methods and Materials

### Study Design, Area, and Study Period

This study employed a cross-sectional design using secondary data from the 2024 Nigeria demographic and health survey, a nationally representative survey that collects comprehensive information on maternal and child health indicators, including immunization coverage. The dataset was accessed from the DHS program https://dhsprogram.com/data/available-datasets.cfm on 4/01/2026 after obtaining the necessary authorization. The NDHS employed a two-stage stratified cluster sampling design to proportionally represent all regions and urban-rural populations. In the first stage, enumeration areas were selected as primary sampling units using probability proportional to size and in the second stage, a fixed number of households per cluster were selected via systematic random sampling. This design ensures national, regional, and subnational representativeness and allows analysis of individual and community level determinants factors while accounting for hierarchical data structure. The survey covered all 36 states and Federal Capital Territory (FCT, Abuja), targeting children aged 12-23 months, their households, and associated maternal and community characteristics. Data were drawn from the kids records (KR) dataset, which includes information on children’s immunization status, maternal and household characteristics, and community level factors. As the study used pre-existing publicly available DHS data, no primary data collection was conducted, and all analysis were performed on the nationally representative dataset provided by the DHS program.

### Study population and sampling

This study focused on children aged 12-23 months, a critical age group for assessing routine immunization coverage. The 2024 NDHS dataset included an unweighted sample of 4,711 children in this age range. Only children with documented DPT1 vaccination status were included, and a child was considered “immunized” if they received at least one dose of DPT1, resulting in a final analytic sample of 4,711 children. The NDHS employed a two-stage stratified cluster sampling design: enumeration areas were selected in the first stage, and households were systematically selected within these enumeration areas in the second stage. All eligible children aged 12-23 months within selected households were included in the study.

### Study variables

#### Dependent variable

The primary outcome of this study was zero-dose immunization, defined as the lack of any routine childhood vaccination, specifically the first dose of the diphtheria-tetanus-pertussis (DPT1) vaccine. This definition is consistent with the WHO classification of zero-dose children, representing those who have not been reached by immunization services. The DPT1 vaccine services as a key indicators of immunization system performance and access to healthcare. Zero-dose status was coded as a binary variable: children who had not received DPT1 were classified as zero-dose (1), while those received at least one dose were classified as vaccinated (0)(8, 10, 22–24). Vaccination status was primarily ascertained from immunization cards, with maternal recall used when cards were unavailable, following survey guidelines for assessing immunization coverage.

#### Independent variable

This study examined both individual and community level factors. Individual level variables included child’s sex, birth order, number of living children, distance from health facilities, maternal or caregiver age, family size, religion, parental education and occupation, family type, marital status, maternal decision making power, household wealth index, place and mode of delivery, number and timing of ANC visiting, postnatal care, and media exposure. Community level variables comprised region of residency classified as north west, north east, north central, south east, south south, south west, and urban versus rural residency.

#### Data analysis

The data were cleaned, recoded and analyzed using STATA version 17.1. Descriptive statistics, including frequency and percentages were used to summarize individual and community level variables.

Sampling weights were applied to account for complex survey design of the demographic and Health survey, including unequal probability of selection and non-response. Variance estimation was then performed using the Taylor series linearization method to produce robust standard errors that appropriately reflect stratification and clustering in survey design.

Given the hierarchical structure of the DHS data, in which children are nested within clusters and observations within the same cluster may be correlated, the assumption of independence of residuals is violated. To account for this clustering effect, a two-level multilevel mixed effect logistic regression model was employed, as traditional single-level models may not adequately address inter-cluster dependencies.

Furthermore, the outcome variable was binary (zero dose), that is vaccinated with at least one dose of DPT1 versus not vaccinated, and therefore a multilevel logistic regression model was appropriate rather than a proportional odds model. Four sequential models were constructed to identify determinants of zero-dose vaccination using Nigerian DHS data. The null model was first fitted, following by model I, which included individual level variables, and model II, which incorporated a community level factors. Finally model III, a mixed-effects model, combined both individual and community level variables.

The intra-cluster correlation coefficient (ICC) was calculated to quantify proportion of total variance attributable-clustering effects or community level variation using the formula ICC=VA/ (VA+3.29), where V_A_ represents community level variance and π^2^/3=3.29 is the variance at individual level for logistic regression.

Model fit was further assessed using the proportional change in variance (PCV), median odds ratio (MOR), and deviance. PCV was computed as 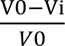, where V0 is the variance in the null model and Vі is the variance in the subsequent models, while MOR was calculated as 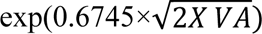. Model III, which included both individual and community level variables, showing the lowest deviance and was considered the best fitting model. Multicollinearity among included variables was checked using spearman’s rank correlation and was not detected. Finally, bivariable logistic regression was performed, and variables with p-values <0.25 were retained for the multivariable multilevel model. In the final model variables with p-values less than 0.05 were considered statistically significantly associated factors of zero-dose children and reported with its adjusted odds ratio (AOR) with 95% confidence interval (CI)

## Result

### Sociodemographic characteristics of the study participants

A total of weighted sample of 4,711 children age 12-23 months and their respondents (women of reproductive age) were included in this study. The children were nearly equally distributed by sex. Most respondents were aged 25-34 years, predominantly resided in rural areas, were married, and belongs to non-polygynous households with varying family sizes. Educational attainment of parents raged from no formal education to higher educations, with a substantial proportion completing at least secondary school. Religiously, participants were mainly affiliated with Islam and Christianity, with a small minority practicing traditional or other religions. Respondents represented diverse socioeconomic background across all wealth quintiles and were drawn from all geographic regions of Nigeria (Table 1).

**Table 1.** Individual sociodemographic characteristics of the study participants and vaccination status among children aged 12-23 months in Nigeria Demographic and health survey 2024.

| Variables | Category | Vaccine status |  |
| --- | --- | --- | --- |
|  |  | Vaccinated with (%) | Unvaccinated with (%) |
| Sex of children | Male | 1,503(62.78) | 891(37.22) |
|  | Female | 1,511(65.21) | 806(34.79) |
| Respondents age | 15-24 | 691(55.02) | 565(44.98) |
|  | 25-29 | 793(64.21) | 442(35.79) |
|  | 30-34 | 789(71.08) | 321(28.92) |
|  | 35-49 | 741(66.76) | 369(33.24) |
| Religion | Catholic | 334(85.9) | 55(14.1) |
|  | Other Christian | 1,173(84.9) | 208(15.1) |
|  | Islam | 1,493(51.25) | 1,420(48.75) |
|  | Traditional and others | 14(50) | 14(50) |
| Residency | Urban | 1,461(79.58) | 375(20.42) |
|  | Rural | 1,553(54.02) | 1,322(45.98) |
| Marital status of caregiver | Never married | 2,731(62.77) | 1,620(37.23) |
|  | Married | 234(83.87) | 45(16.13) |
|  | Previously married | 49(60.49) | 32(39.51) |
| Maternal education | No education | 804(39.90) | 1,211(60.10) |
|  | Primary education | 342(65.77) | 178(34.23 ) |
|  | Secondary education | 1,366(82.64) | 287(17.36) |
|  | Higher education | 502(95.98) | 21(4.02) |
| Father educational status | No education | 649(37.28) | 1,092(62.72) |
|  | Primary | 271(69.13) | 121(30.87) |
|  | Secondary | 1,222(79.51) | 315(20.49) |
|  | Higher | 716(87.42) | 103(12.58) |
| Wealth index | Poorest | 505(41.63) | 708(58.37) |
|  | Poorer | 474(49.84) | 477(50.16) |
|  | Middle | 508(67.44) | 281(32.56) |
|  | Richer | 716(79.12) | 189(20.88) |
|  | Richest | 737(94.61) | 42(5.39) |
| Media exposure | no media exposure | 1,034 (47.7) | 1,135(52.33) |
|  | has media exposure | 1,980(77.89) | 562(22.11) |
| Geographical zone | North west | 609(42.23) | 833(57.77) |
|  | North east | 618(62.68) | 368(37.32) |
|  | North central | 513(61.07) | 327(38.93) |
|  | South east | 460(92.37) | 38(7.63) |
|  | South south | 432(88.16) | 58(11.84) |
|  | South west | 382(83.96) | 73(16.04) |

### Prevalence of zero dose children

A weighted analysis of the 2024 Nigeria demographic and health survey was conducted to estimate the prevalence of zero-dose children aged 12-23 months, accounting for the complex survey design, including clustering, stratification, and sampling weights. The finding indicates that 37.3% (95% CI: 35.1-39.6%) of children in this age group had not received the first dose of the pentavalent vaccine, while 62.7% had received at least one of vaccine.

The prevalence of zero-dos vaccine varied substantially across sociodemographic characteristics, it was highest among children from the poorest household and lowest among those from the richest. Similarly children of mothers with no formal education had substantially higher prevalence compared to those whose mothers attained secondary or higher education. Zero-dose prevalence was also more common in rural areas than in urban settings (Figure 1).

**Figure 1:**
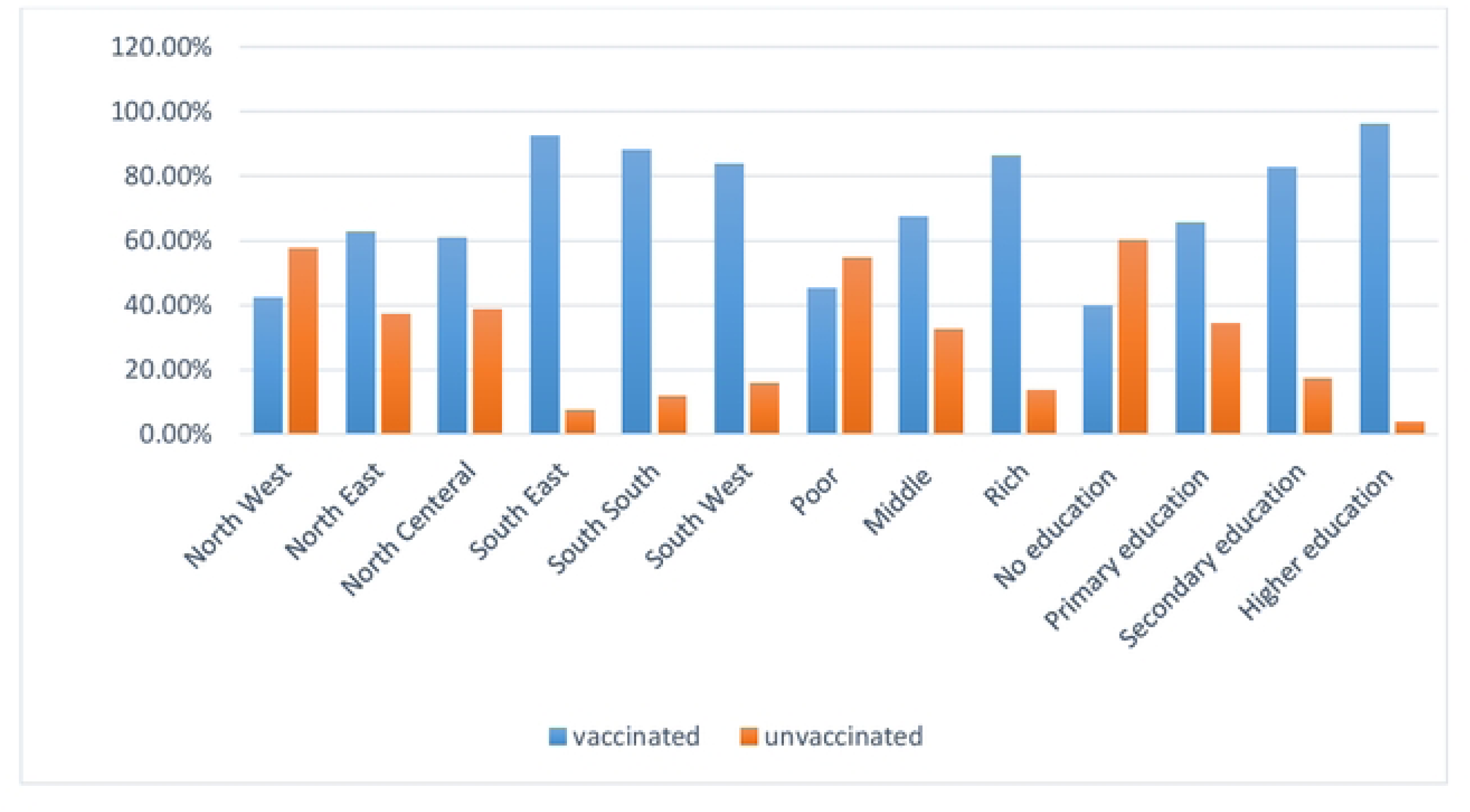
Prevalence of zero-dose vaccination status among children aged 12–23 months by region, wealth index, and maternal education level in Nigeria: analysis of the 2024 DHS Data

### Model fitness and multilevel analysis

To identify the determinants of zero-dose vaccination among children aged 12-23 months in Nigeria, four multilevel model were constructed using the 2024 DHS data. Model performance and fitness were assessed using log-likelihood, deviance, and the Akaike’s information criteria (AIC),while variance components were evaluated through the intra-cluster correlation(ICC), proportional change in variance (PCV), and median odds ratio(MOR) ton account for clustering and community-level effects.

### The random effect (measure of variation) result in multilevel analysis

#### Null model analysis (empty model)

The null model, which contained no explanatory variables, was used to determine the extent of community level clustering, showed a high cluster level variance of 3.73 and an inter cluster correlation (ICC) of 53.1%, indicating that more than half of the variation in zero-dose vaccination was attributable to differences between communities. The MOR was 6.30 reflecting substantial heterogeneity between clusters (Table 2)

**Table 2.** Random effect, measure of variation, and model fitness statistics for zero-dose vaccination across successive models, from null model to the final mixed-effect logistic regression model, among children aged 12-23 months using Nigerian 2024 DHS.

| Parameters | Null model | Model I | Model II | Model III |
| --- | --- | --- | --- | --- |
| Cluster variance ($\sigma^2$) | 3.73 | 0.893 | 2.08 | 0.806 |
| ICC (%) | 53.1 | 21.4 | 38.7 | 19.9 |
| MOR | 6.30 | 2.47 | 3.96 | 2.35 |
| PCV | 0.0% | 76.0% | 44.2% | 78.6% |
| LLR | 836.85, $P<0.001$ | 82.04, $P<0.001$ | 404.52, $P<0.001$ | 69.34, $P<0.001$ |
| Log-likelihood | -2660.426 | -1905 | -2448.546 | -1888.073 |
| Deviance | 5320.85 | 3810 | 4080 | 3776.14 |
| AIC | 5324.85 | 3894 | 4122 | 3856.146 |

Key: ICC inter cluster correlation, MOR median odds ratio, PCV proportional change in variance, AIC Akaike's information criteria, LLK likelihood ratio tests

#### Individual-level model fitness

Inclusion of individual-level variables (model I) substantially improved model fit. The cluster variance decreased to 0.893, ICC declined t0 21.4%, and the MOR fell to 2.47, suggesting that individual characteristics explained a large proportion of the between community variance. The proportional change variance was 76.0%, with a log-likelihood of -1905, deviance of 3810, and AIC of 3894. The likelihood ratio test was significant (X^2^=82.04, p<0.001), confirming the appropriateness of multilevel modeling (Table 2).

#### Community-level model fitness

When community level factors, including geographical zone and place of residency, were added, the model showed a modest reduction in unexplained variance. Cluster variance remained relatively high at 2.08, with an ICC of 38.7% and an MOR of 3.96. The proportional change in variance (PCV) was 44.2%. model fitness indices log-likelihood was -2448.546, deviance was approximately 4080, and AIC was 4122 which were less favorable compared with model I, suggesting that community level factors alone accounted for less variation than individual-level factors (Table 2).

#### Model III (combined or full model)

The final model (model III), incorporating both individual and community level variables, demonstrated that the superior model fitness. The cluster variance decreased significantly to 0.806, with the ICC dropping to 19.9% and the MOR to 2.35, representing the lowest residual heterogeneity between clusters. The proportional change in variance (PCV) attained its maximum values of 78.6% suggesting that the included covariates accounted for most of the observed variation between communities. Modell III also yielded the most favorable fitness statistics, including the lowest log-likelihood (-1888.07), deviance (3776.15), and AIC (3856.15). The significant likelihood ratio test (69.34, p<0.001) confirming that the multilevel model remained a more appropriate fit than a standard single level model (Table 2). Consequently, model III was selected as the final model for reporting and discussion.

### Fixed effect result in multilevel analysis

#### Individual level factors associated with zero-dose vaccination

A multilevel logistic regression model was fitted to examine individual level factors associated with zero-dose vaccination while accounting for clustering at the community level. The analysis showed that birth order, distance to a health facility, maternal age, and occupation, parental education, wealth index, number of ANC visits, postnatal care, place of delivery, and religion were significantly associated factors with zero-dose vaccination among children aged 12-23 months in Nigeria (table 3).

**Table 3.**
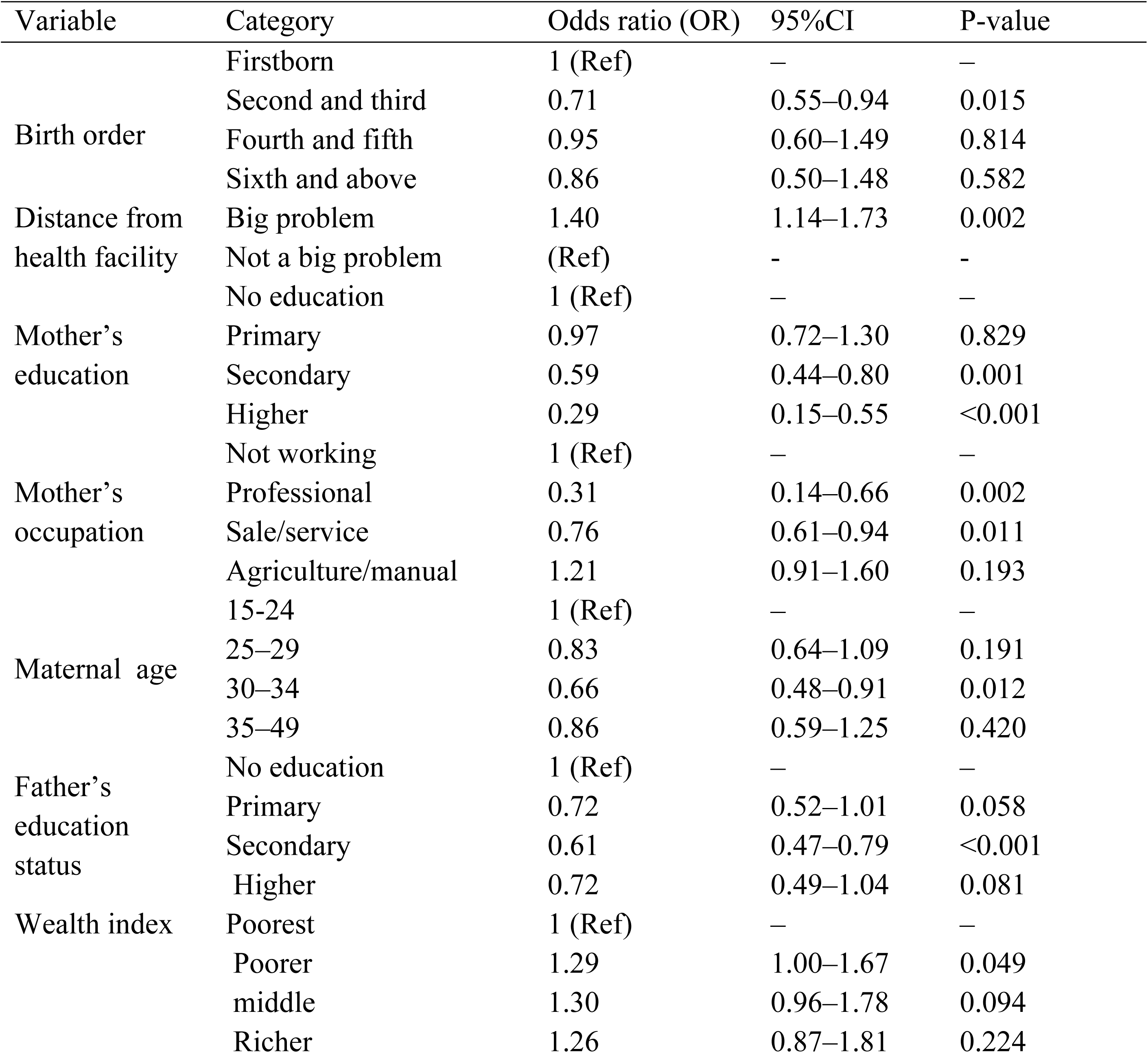
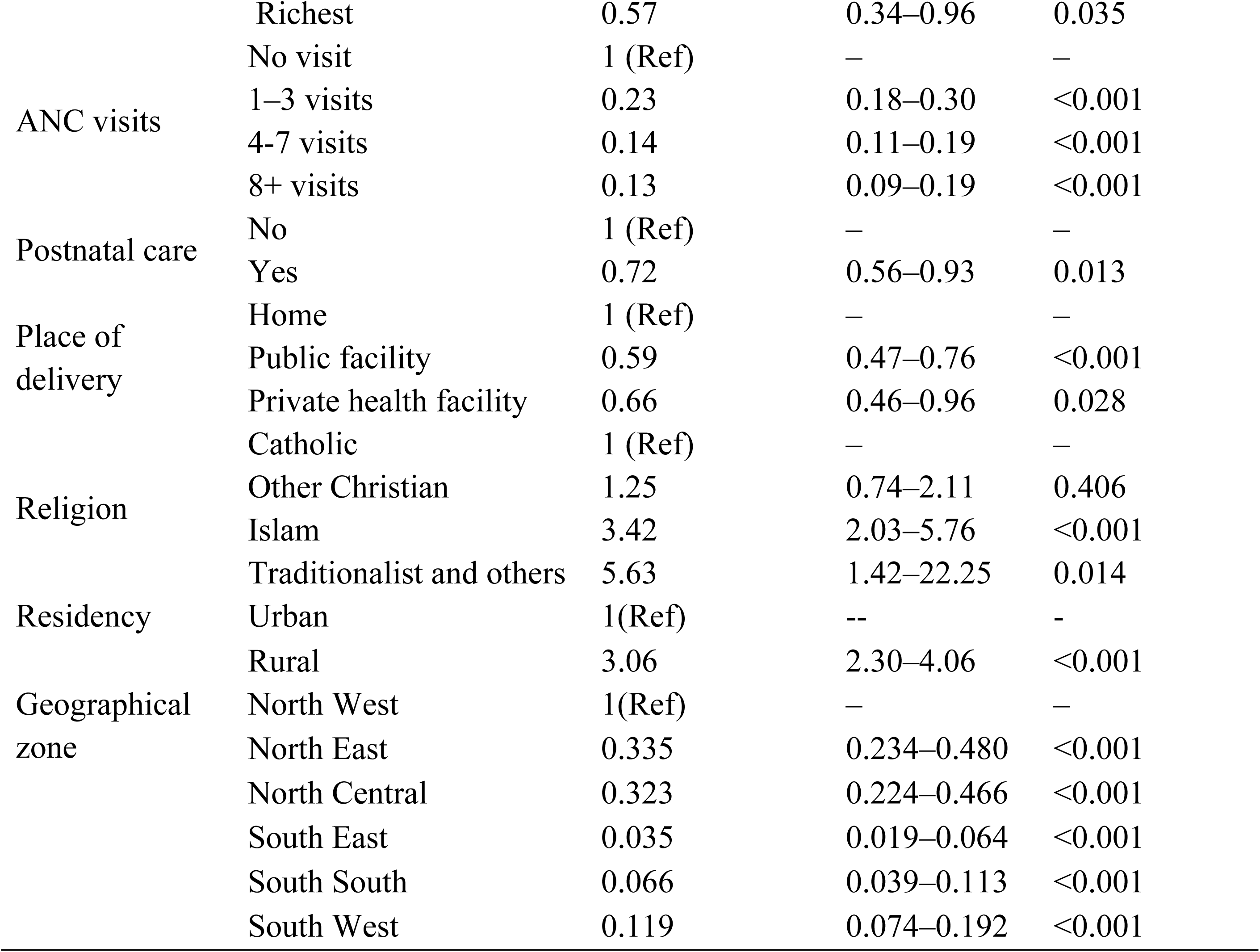
Bivariable logistic regression analysis of individual-and community-level factors associated with zero-dose children aged 12-23 months in Nigeria, 2024 demographic health survey

The mixed effect logistic regression analysis revealed that children of second and third birth order had 29% lower odds of being zero-dose children compared to first-born children (AOR=0.71,95% CI:0.55-0.94). Similarly, maternal age also influenced zero-dose status. Children whose mothers aged 30-34 years were 34% less likely to be zero-dose compared to those whose mothers were 15-24 years old (AOR=0.66; 95% CI:0.48-0.91).

In terms of socioeconomic factors, children from poorer households had 29% higher odds of being zero-dose children compared to those from the poorest households (AOR=1.29; 95% CI:1.00-1.67), whereas children from the richest households were 43% less likely to be zero-dose (AOR=0.57; 95% CI:0.34-0.96). Similarly, maternal occupation showed a significant association with zero-dose status of children. Children whose mothers in professional roles had 69% lower odds of being zero-dose (AOR=0.31; 95% CI: 0.14-0.66), and those whose mothers worked in sales or service sectors had 24% less likely (AOR=0.76; 95% CI: 0.61-0.94) to have be zero-dose, compared with children of not working mothers.

Parental education was a significant determinant of zero-dose children. Children of mothers with secondary education had 41% lower odds of being zero-dose (AOR=0.59; 95% CI: 0.44-0.80), and those whose mothers attained higher education had 71% reduced the chance of to be zero-dose children (AOR=0.29; 95% CI: 0.15-0.55), compared to children of mothers with no formal education. Likewise, children whose fathers had secondary education were 39% less likely to be zero-dose children (AOR=0.61; 95% CI: 0.47-0.79).

Furthermore, access to healthcare services also played a significant role in determinant zero-dose children. Children of mothers reporting distance to a health facility as a major problem had 40% higher chance of being zero-dose children (AOR=1.40; 95% CI: 1.14-1.73). Similarly children born in public health facilities were 41% less likely (AOR=0.59; 95% CI:0.47-0.76), and those born in private facilities were 34% less likely chance of being zero-dose compared to home delivery (AOR=0.66; 95% CI: 0.46-0.96). Additionally, the number of ANC visits was associated with a dose dependent protective effect of zero-dose children. Children of mothers had 1-3 ANC visits had 77% lower odds (AOR=0.23;95% CI: 0.18-0.30), 4-7 visits had 86% more lower odds (AOR=0.14; 95%CI:0.11-0.19), and 8 or more visits had 87% less likely (AOR=0.13,95%CI:0.09-0.19) to be zero-dose children compared with children whose mothers had no ANC visits . Postnatal care further reduced the likelihood of zero-dose vaccination by 28% (AOR=0.72; 95% CI: 0.56-0.93).

Finally, religion was also significantly associated factors of zero-dose vaccination among 12-23 months of children. Children from Muslim religion faith had 3.42 times higher odds of being zero-dose (AOR=3.42; 95% CI: 2.03-5.76), and those practicing traditional or other religion had 5.63 times more likely (AOR=5.63; 95%CI: 1.42-22.25) compared to Christian religion followers, however, fathers occupation, media exposure, sex of child and family type were not significantly associated with zero-dose children among 12-23 months of children in Nigeria.

### Community level factors associated with zero-dose vaccination

A multilevel logistic regression model was fitted to identify the factors of community level factors on zero-dose vaccination, accounting for clustering at the enumeration areas level. Accordingly, rural residency was strongly associated with zero-dose children, with children living in rural communities having more than three times higher odds of being zero-dose children compared to those in urban areas (AOR=3.06; 95% CI:2.30-4.06).

Geographical region was also other strong determinate factors of zero-dose children. Compared to children in the North West, children residing in the north east (AOR=0.33; 95% CI: 0.23-0.48) and north central (AOR=0.32; 95% CI: 0.22-0.47) regions, had approximately 67% and 68% lower odds of being zero-dose children, respectively. More pronounced reduction were observed in the southern regions of the country. South east (AOR=0.04; 95% CI: 0.02-0.06), south south (AOR=0.07; 95% CI: 0.04-0.11), and south west (AOR=0.12; 95% CI: 0.07-0.19) showing substantial lower odds of being zero-dose children (Table3).

### Mixed –effects analysis of factors associated with zero-dose children

The final multivariable multilevel mixed-effects logistic regression model (model III), including 4.711 children across 1, 251 clusters, identified both individual and community-level determinates of zero-dos vaccination among children aged 12-23 months. Significant individual level predictors included maternal and parental education, maternal occupation, birth order, maternal age, household wealth, ANC attendance, place of delivery, postnatal care, distance to the health facilities, and religion. The only community level factors remained significant was geographical region.

At the individual level, birth order was significantly associated with zero-dose vaccination. Children of second or third birth order had 27% lower odds (AOR=0.73; 95% CI: 0.56-0.95), and those of sixth or higher birth order had 35% less likely (AOR=0.65; 95% CI: 0.43-0.97) of being zero-dos compared to first born children. Similarly, children of mothers aged 30-34 years had 36% lower chance of being zero-dose (AOR=0.64; 95% CI: 0.47-0.88) relative to those of younger mothers aged 15-24 years.

Additionally maternal occupation was significant determinant factors of zero-dose vaccination in Nigeria. Children whose mothers worked in professional, occupation was 73% less likely to be zero-dose (AOR=0.27; 95% CI: 0.13-0.58), while those mothers were engaged in sales or services activities were 28% less likely to be zero-dose (AOR=0.72; 95% CI: 0.59-0.89) compared to children of non-working mothers. Likewise, children from the richest households were 56% less likely to be zero-dose vaccination than those from the poorest households (AOR=0.44; 95% CI: 0.26-0.75).

Parental education remained an additional strong predictors of zero-dose vaccination. Comparing with children whose mother had no formal education, those whose mothers attained secondary and higher education had 42% and 72% lower odds of being zero-dose vaccination (AOR=0.58; 95% CI:0.43-0.78) and (AOR=0.28; 95% CI:0.15-0.53) respectively. A similar pattern was observed for parental education, where children of fathers with primary, secondary and higher education had 29%, 41%, and 34% less likely, being zero-dose (AOR=0.71; 95% CI:0.51-0.99), (AOR=0.59; 95% CI:0.45-0.76) and (AOR=0.66; 95% CI:0.46-0.94) respectively, comparing with those whose father had no formal education.

In addition to parental education, healthcare utilization factors demonstrated strong association with zero-dose vaccination. Children of mothers who reported distance to health facilities as a major problem were 42% more likely to be zero-dose than those without such access barriers (AOR=1.42; 95% CI:1.15-1.74). in contrast, children delivered in public health facilities had 35% lower odds of being zero-dos comparing with those delivered at home (AOR=0.65; 95% CI:0.51-0.82). similarly, children whose mother attained 1-3, 4-7 and eight or more ANC visits had 77%,86% and 87% lower odds respectively, of being zero-dose comparing with children whose mothers had no any ANC visits(all p<0.0010). Likewise, receipt postnatal care was associated with a 25% lower likelihood of zero-dose vaccination (AOR=0.75; 95%CI: 0.58-0.97).

In terms of religion, zero-dose vaccination status remained significantly associated with household’s religious affiliation. Comparing with children from catholic and other Christian religion followers, those from Muslim households were 2.58 times more likely to be zero-dose (AOR=2.58; 95% CI:1.47-4.52), while children from traditional or other religious group were five times more likely to remain zero-dose (AOR=5.13; 95% CI:1.31-20.09). At the community level, geographic region also remained a significant predictors of outcome variable. Relative to children residing in north West region, those living in the North East, South East, and South South region had 55%,60%,and 61% lower odds of being zero dose (AOR=0.45; 95% CI:0.33-0.62), (AOR=0.40; 95%CI:0.20-0.82) and (AOR=0.39; 95%CI:0.21-0.73) respectively. However, place of residency (urban versus rural) was not statistically significant in the final model (Table 4).

**Table 4.**
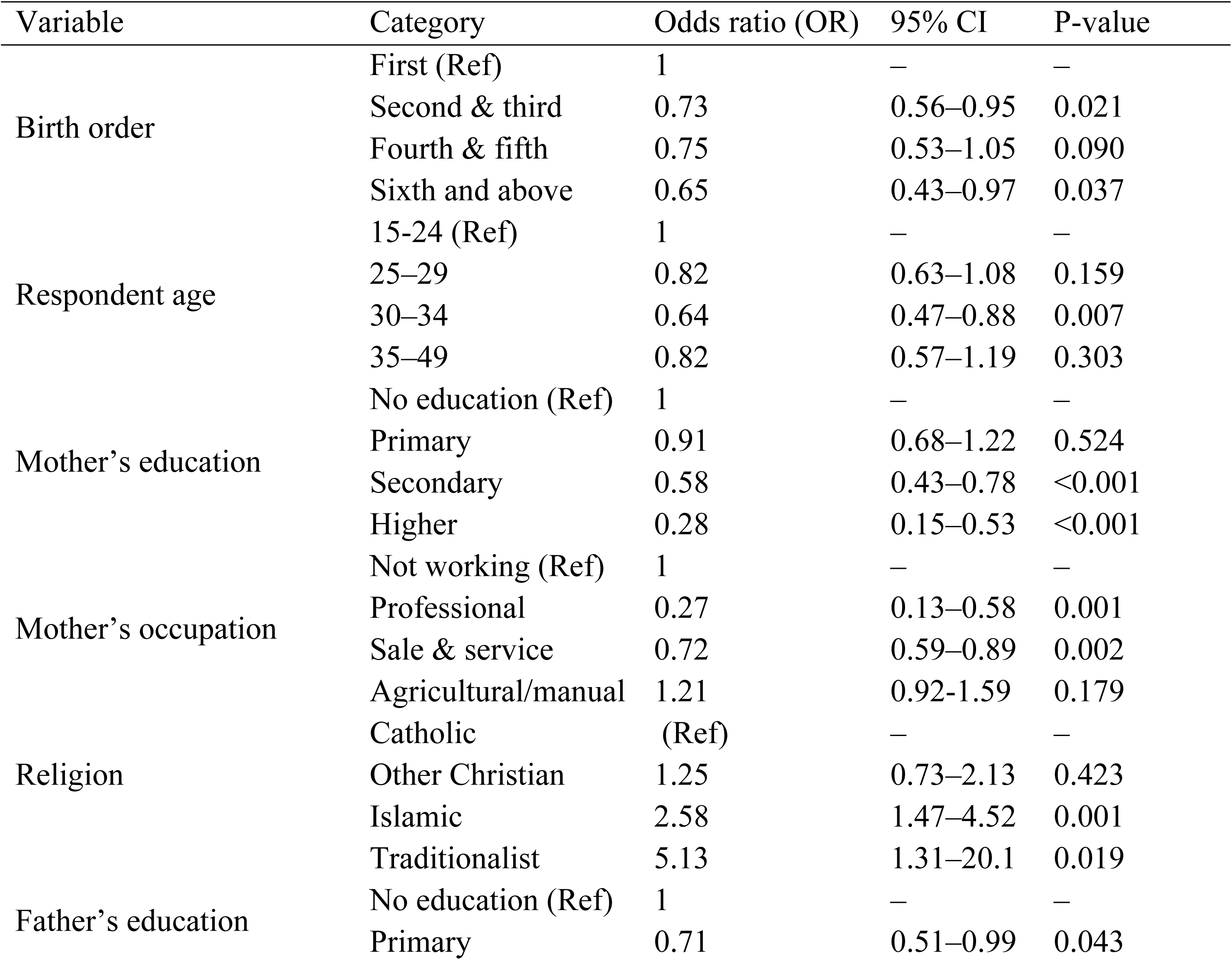
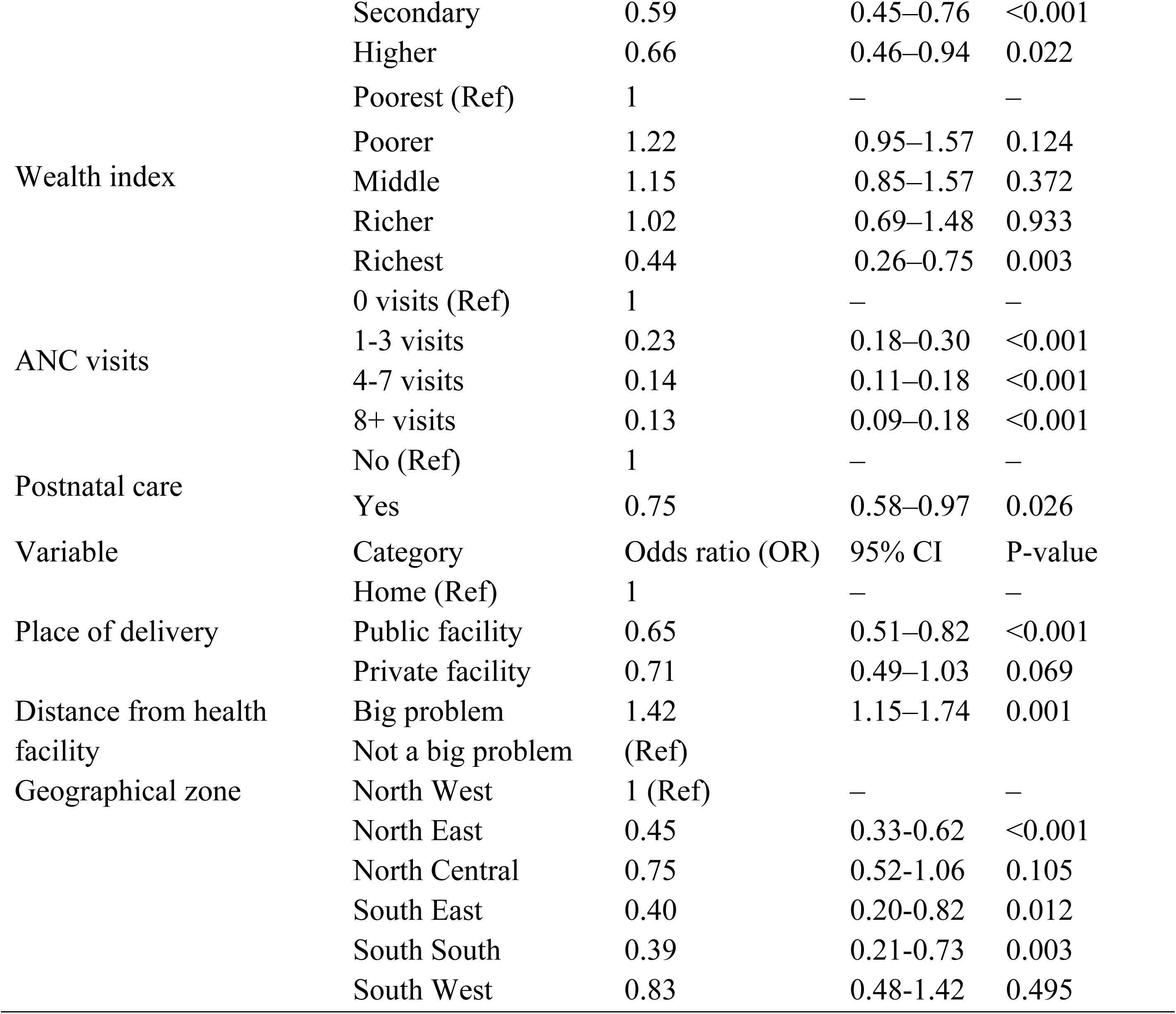
Multilevel mixed-effects logistic regression analysis of determinants of zero-dose vaccination among children aged 12-23 months using Nigerian DHS, 2024.

## Discussion

The present study analyzed data from the Nigeria demographic health survey to examine the determinants of zero-dose children age 12-23 months using a multilevel logistic regression approach. The finding revealed substantial clustering at the community level, as evidenced by a high ICC of 53.1% in the null model, indicating that more than half of the variation in zero-dose status was attributable to differences between communities.

Comparative assessment of model fit statistics identified the full model, which included both individual and community level variables, as the best-fitting model. This model demonstrated the lowest deviance =3776.14 and AIC =3856.146, reflecting improved fit relative to the other models. In addition, it accounted for a large proportion of the between-community variation, with a PCV of 78.6%. The reduction in the MOR from 6.30 in the null model to 2.35 in the final model, along with the decline in ICC to 19.9%, suggests that the included variables substantially explained clustering effects and unobserved heterogeneity across communities. Generally, these findings underscore the importance of both individual and communities-level factors in shaping zero-dose vaccination in Nigeria, as captured in the DHS data.

In this study, birth order was found to be an important factor influencing zero-dose vaccination. Children of higher birth order were less likely to be zero-dose compared with firstborn children. This finding is consistent with evidence from a study conducted in Cameroon (25) which also reported improved vaccination uptake among later-born children. However, it contrasts with findings from pooled analysis across multiple low-and middle-income countries (26–28), where zero-dose prevalence tends to increase with higher birth order and large family size. This discrepancy may reflect contextual differences specific to Nigeria, where factors such as parental experience, childcare practice and interactions with health services may improve with successive births, and potentially enhancing vaccination uptake among later-born children.

The possible justification could be later birth order in this study setting reflects greater maternal familiarity with the health system and strong engagement with vaccination services, resulting in better immunization outcomes compared with firstborns, who may face barriers due to caregiver inexperience. Evidences shows that maternal utilization of health services, including ANC attendance, postnatal care and a positively attitude toward the health system are strong predictors of complete childhood vaccination, suggesting that experienced mothers navigate the system more effectively. Further research on family dynamics, health-seeking behavior, and access to immunization services in Nigeria is needed to clarify the mechanisms underlying the pattern(29, 30).

In this study, both maternal and parental education were significantly associated with the likelihood of zero-dose vaccination among children aged 12-23 months. Children whose mothers had secondary or higher education had significantly lower odds of being zero-dose compared with those whose mother had no formal education. Similarly, children of fathers with secondary or higher were less likely to be zero-dose compared with those whose fathers had no formal education. This aligns with DHS analysis in sub-Saharan Africa (SSA) and other global systematic review and meta-analysis conducted in low-and middle-income countries (31),systematic review and meta-analysis in Ethiopia and DHS data analysis in Nigeria (32–35). The possible reason could be educated parents may have better knowledge and awareness of the benefits and schedules of vaccination, are more likely to engage with health information, and may navigate health services more effectively, all of which facilitate timely vaccination.

The analysis showed that mother’s occupation was significantly associated with zero-dose vaccination status. Compared with children of non-working mothers, those whose mothers were engaged in professional and sales and services worker had 73% and 28% less likely of being zero-dose. In contrast, children of mothers engage in agriculture or manual occupation did not differ significantly from those of non-working mothers in their odds of zero-dose vaccination. This supported by the study from multilevel analysis in Jordan(36), Bangladesh(37),SSA(17), and Ethiopia (33).

These findings suggest that formal employment and economic engagement may enhance access to health services, health information, and social networks that promote immunization uptake. Mothers in professional and services worker are likely to have higher education level, greater health literacy, regular contact with health facilities, and more stable incomes, all of which are known facilitators of vaccination service utilization.

In our study, household wealth status showed a significant association with zero-dose vaccination. Particularly at the highest wealth level. Children from the richest households had 56% lower odds of being zero-dose compared with those from the poorest households, while differences among the poorer, middle, and richer groups were not statistically significant. This findings is consistent with reports from the UNICEF(38),and analysis of DHS data in SSA countries (39), including Nigeria, (35, 40, 41) and Ethiopia (42).

This finding can be explained by the advantages enjoyed by wealthier households, including better access to healthcare services, higher health awareness, and greater ability to overcome indirect cost such as transportation and time. They are also more likely to reside in urban areas and maintain regular contact with health providers, increase opportunities for timely vaccination. In contrast, the absence of significant difference among poorer, middle, and richer groups may reflect shared structural barriers or the partial effectiveness of equity-focused immunization program generally, this suggest that the protective effect of wealth is most pronounced at the highest socioeconomic level, rather than following a gradual gradient across all wealth quintiles.

Our research finding demonstrate a significant association between ANC attendance and zero-dose vaccination. Compared with mothers who had no ANC visits, those with higher ANC attendance were significantly less likely to have zero-dose children. Similar result have been reported by the WHO and DHS analysis of 47 low-and middle-income countries which demonstrated that women attending four or more ANC visits have significantly higher odds of completing their children immunization compared with those who have no ANC visits(43–47).

This inverse association could be due to ANC visits offer mothers critical opportunities for education, counseling, and linkage preventive services. This all enhance maternal knowledge, health literacy, and trust in the health care system, promoting timely uptake of vaccine and reducing the likelihood of zero-dose children. Conversely, mothers who do not attend ANC are less likely to receive information regarding immunization, face greater access barriers, and are at higher risk of leaving their children unvaccinated.

Similarly, children delivered in public health facilities had significantly lower odds of being zero-dose compared to those home delivery. This aligned with previous WHO, UNICEF report(8, 48), global evidence conducted by START center (22), scoping review in developed countries(49) and studies from SSA(21, 39, 50), which consistently showed that institutional delivery is positively associated with lower zero-dose vaccination prevalence.

Additionally, children whose caregiver’s reported distance as a big problem had higher odds of being zero-dose. This is consistent with a global evidence analysis by the START Center (22), scoping review in developed countries(49), and large body of literature across low-and middle – income countries (39, 50, 51). This is could be due to longer travel time, poor road infrastructure, and lack of transportation significantly reduce the likelihood of accessing immunization services.

Furthermore, religion was found to be a significant determinant of zero-dose vaccination among children age 12-23 months. Children from Islamic households had 2.58 times higher odds of being zero-dose, whereas children from traditionalist households exhibited the highest likelihood of being unvaccinated. Similar pattern have been documented across 66 low-and middle-income countries, SSA and India (22, 52–54). This could be explained by the influence of religious beliefs and cultural norms on caregiver’s attitude towards immunization. Such factors can shape perception of vaccine safety and effectiveness, affect level of trust in the health system, and influence the acceptability of modern health interventions. This beliefs may leads caregivers to prioritize health practice over formal immunization services.

The present study demonstrates that geographic zone within Nigeria is a significant determinant of zero-dose vaccination status among children aged 12-23 months. Children residing in the north east, south east, and south south zones had significantly lower odds of being zero-dose, while no statistically significant difference was observed for those living in the north central zone compared with their counterparts in the south west.

This regional disparities may be attributed to variations in health system capacity and infrastructure, level of maternal education, access to immunization services, and underling cultural factors. Collectively, these findings underscore the importance of geographically targeted, context specific interventions to effectively reach under-vaccinated population in Nigeria.

### Conclusion and recommendation

This multilevel analysis of the 2024 Nigeria Demographic and Health Survey reveals that zero-dose vaccination remains a critical public health concern in Nigeria, with a substantial proportion of children age 12-23 months not receiving even the first dose of a DPT containing vaccine. The burden of zero-dose vaccination is disproportionately concentrated among children from socioeconomically disadvantaged households, those born to mothers with low educational attainment, and those living in underserved geographic regions.

Both individual-level and community-level factors significantly influence of zero-dose status, including maternal and parental education, household wealth status, maternal employment, ANC attendance, place of delivery, postnatal care utilization, religion, perceived distance to health facilities and geographical region, with marked regional disparities.

Generally, these findings highlight the need for equity-focused interventions that strengthen maternal health service utilization, improve immunization coverage across the continuum of care, and address socioeconomic, religious, and regional inequalities to reduce zero-dose prevalence in Nigeria

## Data Availability

The data underlying this study are third-party data from the Demographic and Health Survey (DHS) Program and are subject to legal and ethical access restrictions imposed by the data owner. The authors did not have special access privileges. Researchers can obtain the data by requesting access through the DHS Program website (https://dhsprogram.com/data/) after registration and approval.

## Abbreviations

AOR: Adjusted Odd ratio
AIC: Akaike’s information criteria
ANC: Antenatal care
CI: Confidence Interval
DHS: Demographic health survey
DPT1: Diphtheria-tetanus-pertussis
DPT1cv1: Diphtheria-tetanus-pertussis-containing vaccine
EPI: Expanded program on Immunization
IA2030: Immunization Agenda 2030
ICC: Intra-cluster correlation coefficient
LLR: Likelihood ratio test
MOR: Median odds ratio
PCV: Proportional change in variance
SSA: Sub-Saharan Africa
UNICEF: United Nations International Children’s Emergency Fund
WHO: World Health Organization.

## Declarations

## Ethics approval and consent to participant

As this was a secondary data analysis of EDHS, informed consent from participants was not required. Data requests and consent for access were acquired from DHS International. All data was anonymized before obtaining informed consent from DHS International.

## Consent for publication

Not applicable

### Data availability

All data generated and analyzed is provided with in manuscript or as supplementary file

### Clinical trial

### Not applicable

#### Funding

No funding is obtained for the this review system

### Authors’ contribution

Conceptualization: DM. Data curation: DM. Formal analysis: DM, AS. Investigation: DM, AS, Methodology: DM, TK. Project administration: DM, TK. Software: DM, AS, JM, Supervision: AD. Visualization: DM, AS. Writing of original draft: DM, AS. Writing review and editing: DM, AS, TK, AD, TL, JM,

## Acknowledgement

We would like to acknowledge DHS international to access this data

